# Clinical characteristics and reasons of different duration from onset to release from quarantine for patients with COVID-19 Outside Hubei province, China

**DOI:** 10.1101/2020.03.21.20038778

**Authors:** Suochen Tian, Zhenqin Chang, Yunxia Wang, Min Wu, Wenming Zhang, Guijie Zhou, Xiuli Zou, Hui Tian, Tingfang Xiao, Junmin Xing, Juan Chen, Jian Han, Kang Ning, Tiejun Wu

**Affiliations:** Department of Intensive Care Unit, Liaocheng people’s hospital, Liaocheng, Shandong, 252000, China; Department of Hemodialysis, Liaocheng people’s hospital, Liaocheng, Shandong, 252000, China; Department of Intensive Care Unit, Liaocheng Infectious Diseases Hospital, Liaocheng, Shandong, 252000, China; Department of Nosocomial Infection, Liaocheng people’s hospital, Liaocheng, Shandong, 252000, China; Department of Respiratory Medicine, Liaocheng people’s hospital, Liaocheng, Shandong, 252000, China; Department of Pulmonology, The Affiliated Hospital of Shandong University of TCM, Jinan, Shandong, 250000, China; Department of Respiratory Medicine, The First Affiliated Hospital of Shandong First Medical University, Jinan, Shandong, 250000, China

**Keywords:** COVID-19 patients, quarantine, characteristic

## Abstract

**Objective:** To find out more characteristics and rules of COVID-19 by analyzing the clinical course of COVID-19 patients in a region outside Hubei province.

**Methods:** 37 cases diagnosed adult COVID-19 cases of general characteristics, history of epidemiology, chronic underlying diseases, clinical symptoms and complications, chest CT, biochemical monitoring, severity assessment, treatment and outcome were retrospectively analyzed, and according to the duration from onset to release from quarantine were divided into ≤20 -day group and > 20 -day group, compare the similarities and differences between the two groups.

**Results:** Among the 37 patients, 5 were mild, 30 were moderate, 1 was severe and 1 was critical. All the patients were released from quarantine without death. The average duration from onset to release from quarantine was 20.2±6.6 days, The average length of stay from onset to hospitalization was 4.1±3.7 days, and hospitalization duration average 16.1 ±6.2 days. The average age was 44.3±1.67 years. 78.4% of cases were caused by exposure to a confirmed patient or the workplace of a confirmed patient. The main symptoms were cough (67.6%), fever (62.2%), shortness of breath (32.4%), fatigue (24.3%), sore throat (21.6%,) vomiting or diarrhea (21.6%). The white blood cell count was decreased in 27.0% of the patients, and the lymphocyte count was decreased in 62.2% of the patients, of which 43.5% patients were ≤0.6×109/L. On admission, 86.5% of patients with chest CT showed pneumonia, including some asymptomatic patients. 68.8% of patients showed bilateral infiltration. In the > 20-day group, the average age was 49.9±1.38 years old, and the duration from onset to hospitalization was 5.5±3.9 days. Compared with the ≤ 20-day group, the age was older and duration was longer, P < 0.05. All the 7 asymptomatic patients were ≤20 –day group. When 37 patients were released from quarantine, the white blood cell count of 16.2% patients was < 4.0×10^9/L^, and the lymphocyte count of 59.5% patients was <1.1×10^9/L^, and the absolute count of white blood cells and lymphocytes was 5.02±1.3 4×10^9/L^ and 1.03±0.34×10^9/L^ respectively, compared with those on admission, P > 0.05.

**Conclusion:** The majority of COVID-19 cases in the study area were mild and moderate, with good clinical outcomes. There were some special characteristics in the clinical process. The reasons of duration from onset to release from quarantine were complex. There was no significant change in the number of granulocytes at the time of release from quarantine compared to the time of admission.

## Background

Previous articles have introduced the clinical characteristics and outcomes of COVID-19 ^[1,2,3,4,5,6]^. These were mainly early cases in Hubei province, especially in Wuhan, and due to the limitation of insufficient medical conditions at that time, this had a certain impact on the outcome and treatment of COVID 19. The clinical characteristics and outcomes of patients in Hubei province and outside of Hubei province were different ^[1]^. There was an article on the early clinical characteristics of 13 COVID - 19 patients outside Hubei province, but the number was too small and the article only described the early clinical characteristics ^[7]^. Liaocheng city, in the Middle East of China, is a prefecture-level city in Shandong province with a population of more than 6 million. As a region outside Hubei province, what are the similarities and differences between the characteristics of the cases here and those of Hubei province and even other countries and regions? Besides, are there any special rules for patients who cannot be released from quarantine for a long time?

## Methods

The diagnosis, release from quarantine and severity of all cases were determined in accordance with the protocol of diagnosis and treatment of novel coronavirus pneumonia issued by National Health Commission of the People’s Republic of China and National Administration of Traditional Chinese Medicine ^[8,9]^. A confirmed case was defined as a positive result to real-time reverse-transcriptase polymerase-chain-reaction (RT-PCR) assay for nasal and pharyngeal swab specimens^[10]^.

The criteria for all cases to be released from quarantine were as recommended in the above protocol, starting with the following three:

1. Body temperature returns to normal for more than 3 days; 2. Respiratory symptoms improved significantly; 3. Pulmonary imaging showed significant absorption of acute exudative lesions. On this basis, strict nucleic acid negative test standard: after 5 days in hospital, every 2 days test, to nucleic acid test negative case, instead of every 24 hours test, three consecutive test negative, they can be released from quarantine. During the dynamic test, if nucleic acid negative cases were once again positive or suspicious, the above steps were restarted. Some cases were kept in the hospital for 14 days after they were released from quarantine.

Severe cases were identified in accordance with respiratory criteria, excluding those who did not meet respiratory criteria and require intensive management ^[11]^.

Of the 38 patients, one was 5 months old, a patient identified by screening and released from quarantine after 9 days in hospital, and was excluded from this analysis. The other 37 patients were all adults. For the incubation period, considering the characteristics of the patients in this study, the time of the first infection with the novel coronavirus was difficult to be precise, so it was not discussed.

This study retrospectively analyzed the general characteristics, epidemiological history, chronic underlying diseases, clinical symptoms and complications, chest CT, biochemical monitoring, severity assessment, treatment plan and outcome of 37 patients. The results of the examination at the study time nodes were reported as ±24 hours. 1 patient lacked serum amyloid A (SAA) and c-reactive protein (CRP) on admission, and 1 patient lacked erythrocyte sedimentation rate (ESR) on admission, which were excluded from the corresponding item statistics. In addition, these patients were divided into ≤20-day group and > 20-day group according the duration of release from quarantine, and the similarities and differences between the two groups in the clinical process were compared, in order to find the relevant factors of nucleic acid continued positive.

## Statistical analysis

Continuous variables were expressed as the means and standard deviations or medians and interquartile ranges (IQR) as appropriate. Categorical variables were summarized as the counts and percentages in each category. T test or Wilcoxon rank-sum tests were applied to continuous variables, and Fisher’s exact tests were used for categorical variables, and Wilcoxon rank sum test was used for rank classification variables. All analyses were conducted with SPSS software version 23.0.

## Results

The duration from onset to admission of 37 confirmed patients ranged from 1 day to 10 days, and the number of asymptomatic patients detected by screening was calculated as 1 day. The shortest length of stay was 7 days, the longest was 32 days. From onset to release of quarantine, the shortest was 8 days, the longest was 34 days, and it was 29 days for 1 severe patient and 11 days for 1 critical patient.

According to the pneumonia severity index (PSI) on admission, 89.2% (17+16/37) of patients were classified as low risk grade I and II. In terms of epidemiological history, all the six patients initially diagnosed had a history of sojourn in Wuhan, and the subsequent patients were mainly in contact with the confirmed patients or their workplaces. High rates of symptoms on admission were cough, fever, shortness of breath, fatigue, sore throat, vomiting or diarrhea, and 87.0% of the patients had low fever. No abnormalities of platelets, creatine kinase, or creatinine were observed in the routine blood biochemistry on admission (table I). During the treatment, 2 patients with acute respiratory distress syndrome (ARDS), 1 with mild and 1 with moderate ^[12]^, were given high-flow nasal cannula (HFNC) oxygen therapy without noninvasive or invasive mechanical ventilation. Plasma exchange was performed in critically ill patients with moderate ARDS. No serious complications such as shock, kidney injury, pulmonary embolism, and diffuse intravascular coagulation occurred in all patients. In the treatment, 1 critically ill patient received an antifungal drug, and all patients received two or more antiviral drugs. The application ratio of other therapeutic measures was in order: thymosin, oxygen therapy, albumin, hormone, immunoglobulin. The application proportion of Traditional Chinese medicine (TCM) was 100%, including Chinese medicine preparation, acupuncture, moxibustion, etc. (table II).

**Table 1.** Clinical Characteristics of the Study Patients on Admission

| Characteristics | All Patients<br>(n=37) | Duration from Onset to Release from Quarantine |  |  |
| --- | --- | --- | --- | --- |
|  |  | ≤20-day | >20-day | P |
|  |  | (n=19) | (n=18) |  |
| Age (yr) | 44.3±1.67 | 39.1±1.79 | 49.9±1.38 | 0.047 |
| Gender |  |  |  |  |
| Male (%) | 17/37 (45.9) | 9/37(47.4) | 8/37(44.4) | 0.121 |
| Female (%) | 20/37 (54.1) | 10/37(52.6) | 10/37(55.6) |  |
| Duration from onset to hospitalization(d) | 3(5.5) | 2(4) | 6(5.25) | 0.016 |
| PSI | 45(105) | 51(53.5) | 44.5(55.75) | 0.932 |
| I (%) | 17/37(40.5) | 8/19(42.1) | 9/18(38.9) |  |
| II≤70 (%) | 16/37(43.2) | 10/19(52.6) | 6/18(33.3) |  |
| III71-90 (%) | 3/37(8.1) | 1/19(5.3) | 2/18(11.1) |  |
| IV91-130 (%) | 1/37(2.7) | 0/19(0) | 1/18(5.6) |  |
| V>130 (%) | 0/37(0) | 0/19(0) | 0/18(0) |  |
| Underlying disease |  |  |  |  |
| Any(%) | 8/37(21.6) | 4/19(21.1) | 4/18(22.2) | 1 |
| Hypertension (%) | 3/37(8.1) | 1/19(5.3) | 2/18(11.1) | 0.604 |
| Coronary heart disease (%) | 3/37(8.1) | 1/19(5.3) | 2/18(11.1) | 0.604 |
| Diabetes (%) | 2/37(5.4) | 1/19(5.3) | 1/18(5.6) | 1 |
| Pulmonary interstitial fibrosis (%) | 1/37(2.7) | 0/19(0) | 1/18(5.6) | 0.486 |
| Cirrhosis, Liver cancer (%) | 1/37(2.7) | 0/19(0) | 1/18(5.6) | 0.486 |
| Epidemiological history |  |  |  |  |
| Contact with wild (%) | 0/37(0) | -- | -- |  |
| Wuhan sojourn (%) | 6/37(16.2) | 3/19(15.8) | 3/18(16.7) | 1 |
| Contact with a diagnosed patient or workplace (%) | 29/37(78.4) | 15/19(78.9) | 14/18(77.7) | 1 |
| Not clear (%) | 2/37(5.4) | 1/19(5.3) | 1/18(5.6) | 1 |
| Symptoms |  |  |  |  |
| Asymptomatic (%) | 7/37(18.9) | 7/19(36.8) | 0/18(0) | 0.008 |
| Fever (%) | 23/37(62.2) | 8/19(42.1) | 15/18(83.3) | 1 |
| ≤38.0℃ (%) | 20(87.0) | 7(87.5) | 13(86.7) | 1 |
| >38.0℃ (%) | 3(13.0) | 1(12.5) | 2(13.3) | -- |
| Chills (%) | 1/37(2.7) | 0/19(0) | 1/18(5.6) | 0.486 |
| Fatigue (%) | 9/37(24.3) | 1/19(5.3) | 8/18(44.4) | 0.008 |
| Headache (%) | 3/37(8.1) | 0/19(0) | 3/18(16.7) | 0.105 |
| Nasal congestion (%) | 3/37(8.1) | 2/19(10.5) | 1/18(5.6) | 1 |
| Sore throat (%) | 8/37(21.6) | 0/19(0) | 8/18(44.4) | 0.001 |
| Cough (%) | 25/37(67.6) | 11/19(57.9) | 14/18(77.8) | 0.295 |
| Hemoptysis (%) | 1/37(2.7) | 0/19(0) | 1/18(5.6) | 0.486 |
| Shortness of breath (%) | 12/37(32.4) | 3/19(15.8) | 9/18(50.0) | 0.035 |
| Vomiting or diarrhea (%) | 8/37(21.6) | 5/19(26.3) | 3/18(16.7) | 0.693 |
| Pain in a muscle or joint (%) | 2/37(5.4) | 1/19(5.3) | 1/18(5.6) | 1 |
| Laboratory findings |  |  |  |  |
| WBC<4.0×10<sup>9</sup>/L (%) | 10/37(27.0) | 4/19(21.1) | 6/18(33.3) | 1 |
| L<1.1×10<sup>9</sup>/L (%) | 23/37(62.2) | 12/19(63.2) | 11/18(61.1) | 0.366 |
| L≤0.6×10<sup>9</sup>/L (%) | 10/23(43.5) | 6/12(50.0) | 4/11(36.4) | 0.68 |
| SAA>10mg/L (%) | 21/37(56.8) | 10/19(52.6) | 11/18(61.1) | 0.743 |
| CRP>5mg/L (%) | 19/37(51.4) | 7/19(36.8) | 12/18(66.7) | 0.103 |
| PCT>0.5ng/ml (%) | 1/37(2.7) | 0/19(0) | 1/18(5.6) | 0.486 |
| LDH>250u/L (%) | 6/37(16.2) | 1/19(5.3) | 5/18(27.8) | 0.09 |
| Alb<40g/L (%) | 24/37(64.9) | 9/19(47.4) | 15/18(83.3) | 0.038 |
| AST>40u/L (%) | 4/37(10.8) | 3/19(15.8) | 1/18(5.6) | 0.604 |
| ALT>40u/L (%) | 2/37(5.4) | 1/19(5.3) | 1/18(5.6) | 1 |
| TBL>17.1ummol/L (%) | 13/37(35.1) | 5/19(26.3) | 8/18(44.4) | 0.313 |
| D -dimer >0.5mg/L (%) | 7/37(18.9) | 4/19(21.1) | 3/18(16.7) | 1 |
| Fib>4g/L (%) | 9/37(24.3) | 4/19(21.1) | 5/18(27.8) | 0.714 |
| ESR≥20mm/h (%) | 24/37(64.9) | 10/19(52.6) | 14/18(77.8) | 0.17 |
| Chest CT |  |  |  |  |
| Pneumonia (%) | 32/37(86.5) | 16/19(84.2) | 16/18(88.9) | 1 |
| Bilateral infiltration (%) | 22(68.8) | 9(56.2) | 13(81.2) | 0.252 |
| Unilateral infiltration (%) | 10(31.2) | 7(43.8) | 3(18.8) |  |

PSI: Pneumonia Severity Index; WBC: White Blood Cells; L: Lymphocyte; SAA: Serum Amyloid
A; CRP: C-Reactive Protein; PCT: Procalcitonin; LDH: Lactate Dehydrogenase; Alb: Albumin;
AST: Aspartate Aminotransferase; ALT: Alanine Aminotransferase; TBL: Total Bilirubin; Fib:
Fibrinogen; ESR: Erythrocyte Sedimentation Rate.

**Table II.** Severity Classification of Patients and Complications and Treatment Measures before Release form Quarantine

| Characteristics | All Patients<br>(n=37) | Duration from Onset to Release from Quarantine |  |  |
| --- | --- | --- | --- | --- |
|  |  | ≤20-day<br>(n=19) | >20-day<br>(n=18) | P |
| Severity |  |  |  | 0.738 |
| Mild (%) | 5(13.5) | 3(15.8) | 2(11.1) |  |
| Moderate (%) | 30(81.1) | 15(78.9) | 15(83.3) |  |
| Severe (%) | 1(2.7) | 0(0) | 1(5.6) |  |
| Critical (%) | 1(2.7) | 1(5.3) | 0(0) |  |
| Complications |  |  |  | 1 |
| ARDS (%) | 2 (5.4) | 1(5.3) | 1(5.6) |  |
| Treatment |  |  |  |  |
| Antibiotics | 27 (73) | 12 (63.2) | 15 (83.3) | 0.269 |
| Intravenous antibiotics (%) | 17/27(63.0) | 9/12(75.0) | 8/15(53.3) | 0.424 |
| Oral antibiotics (%) | 10/27(37.0) | 3/12(25.0) | 7/15(46.7) |  |
| Antifungal drugs | 1/37(2.7) | 0/19(0) | 1/18(5.6) | -- |
| Antiviral drugs (%) | 37/37(100) | 19/19(100) | 18/18(100) | 0.17 |
| Two (%) | 25(67.6) | 15(78.9) | 10(55.6) |  |
| Three (%) | 12(32.4) | 4(21.1) | 8(44.4) |  |
| Glucocorticoids (%) | 8(21.6) | 4(21.1) | 4(22.2) | 1 |
| Daily dose |  |  |  | 0.131 |
| 40mg (%) | 6/8(75) | 4/4(100) | 2/4(50.0) |  |
| 80mg (%) | 1/8(12.5) | 0/4(0) | 1/4(25.0) |  |
| 120mg (%) | 1/8(12.5) | 0/4(0) | 1/4(25.0) |  |
| Albumin (%) | 12(32.4) | 6(31.6) | 6(33.3) | 1 |
| Immunoglobulin (%) | 7(18.9) | 2(10.5) | 5(27.8) | 0.232 |
| Thymosin (%) | 24/37(64.9) | 9/19(47.4) | 15/18(83.3) | 0.038 |
| Oxygen therapy (%) | 15/37(40.5) | 7/19(36.8) | 8/18(44.4) | 0.743 |
| Common (%) | 13/15(86.7) | 6/7(85.7) | 7/8(87.5) | 1 |
| HFNC (%) | 2/15(13.3) | 1/7(14.3) | 1/8(12.5) | -- |
| PE (%) | 1/37(2.7) | 0/19(0) | 1/18(5.6) | 0.486 |
| TCM (%) | 37/37(100) | 19/19(100) | 18/18(100) | -- |

HFNC: High-Flow Nasal Cannula; PE: Plasma Exchange; TCM: Traditional Chinese Medicine.

In the >20-day group, the age was older and the length of stay from onset to hospitalization was longer (P>0.05). As for the clinical symptoms on admission, all the 7 asymptomatic patients were ≤20 -day group. Symptoms of fatigue, sore throat, and shortness of breath were higher in the >20-day group (table I), and patients were treated with more albumin and thymus peptide (table II), and had longer hospital stays (table III).

**Table III.** Clinical Outcomes of Patients

| Outcomes | All Patients<br>(n=37) | Duration from Onset to Release from Quarantine |  | P |
| --- | --- | --- | --- | --- |
|  |  | ≤20-day<br>(n=19) | >20-day<br>(n=18) |  |
| Hospitalization duration (d) | 16.1±6.2 | 12.4±3.7 | 20.1±5.8 | 0 |
| Duration from onset to release from quarantine (d) | 20.2±6.6 | 15.1±3.4 | 25.6±4.4 | 0 |
| Blood findings at the time of release from quarantine |  |  |  |  |
| WBC <4.0×10<sup>9</sup>/L (%) | 6/37(16.2) | 2/19(10.5) | 4/18(22.2) | 0.405 |
| L <1.1×10<sup>9</sup>/L (%) | 22/37(59.5) | 10/19(52.6) | 12/18(66.7) | 0.508 |
| L ≤0.6×10<sup>9</sup>/L (%) | 4/22(18.2) | 2/10(20.0) | 2/12(16.7) | 1 |
| SAA>10mg/L ( % ) | 5/37(13.5) | 3/19(15.8) | 2/18(11.1) | 1 |

**Table IV.** Granulocyte Counts on Admission and Release from Quarantine

| Granulocyte counts | All patients (n=37) |  |  | ≤20-day(n=19) |  |  | > 20-day(n=18) |  |  |
| --- | --- | --- | --- | --- | --- | --- | --- | --- | --- |
|  | Admission | Release | P | Admission | Release | P | Admission | Release | P |
| WBC (×10<sup>9</sup>/L) | 5.26±2.15 | 5.02±1.34 | 0.56 | 5.95±2.38 | 5.33±1.30 | 0.33 | 4.54±1.64 | 4.67±1.34 | 0.8 |
| L (×10<sup>9</sup>/L) | 0.94(0.73) | 1.05(0.58) | 0.599 | 0.8(0.76) | 1(0.6) | 0.5 | 1.1(0.74) | 1.1(0.31) | 0.894 |
| WBC<4.0×10<sup>9</sup>/L ( % ) | 10/37(27.0) | 6/37(16.2) | 0.398 | 4/19(21.1) | 2/19(10.5) | 0.66 | 6/18(33.3) | 4/18(22.2) | 0.715 |
| L <1.1×10<sup>9</sup>/L ( % ) | 23/37(62.2) | 22/37(59.5) | 1 | 12/19(63.2) | 10/19(52.6) | 0.743 | 11/18(61.1) | 12/18(66.7) | 1 |
| L ≤0.6×10<sup>9</sup>/L ( % ) | 10/23(43.5) | 4/22(18.2) | 0.108 | 6/12(50.0) | 2/10(20.0) | 0.204 | 4/11(36.4) | 2/12(16.7) | 0.371 |

## Discuss

The small number of cases in this study affected the statistical analysis of some of the results, but it still showed many obvious characteristics.

The 37 patients were mainly mild and moderate, with only 1 severe case and 1 critical case. Finally, all patients were released from quarantine without death, and the clinical outcome was significantly better than that in Hubei province. ^[1,5,6,13]^ The main reason may be that the patient was relatively mild and the availability of adequate medical facilities and personnel made the patient less likely to develop into a severe or critical condition. After hospitalization, all patients were stratified according to the PSI, which may be a better strategy to improve the outcome of COVID-19 patients, especially in the outbreak where medical resources are relatively insufficient ^[14]^. Although the proportion of patients with chronic underlying diseases in this group was 21.6%, it did not seriously affect the outcome, since most of the patients were around 44 years old. According to the epidemiological history, some of the patients in this study were in direct contact with the confirmed patients, and some were in the same workplace and did not meet the conceptual standard of close contact, which suggested the existence of the transmission route of COVID -19 aerosol. Among these patients, the proportion of asymptomatic patients accounted for 18.9% of the total patients. In some patients, chest CT scan still showed pneumonia and decreased white blood cell and lymphocyte counts. Among the first few symptoms on admission, the proportion of shortness of breath and gastrointestinal symptoms was high, which could not exclude the influence of psychological factors. In addition, the proportion of patients with fever was not high, and the fever was mainly low. These characteristics were different from other highly infectious respiratory virus infections.^[15,16]^Among the routine blood biochemistry on admission, the most common was a reduction in the lymphocyte count, and in nearly half of these patients, the number was ≤ 0.6×10^9/L^. White blood cell counts were mostly normal, and less than a third of patients were lower. Other abnormalities with relatively high proportions were ESR, albumin, SAA and CRP, but most of the changes were slight and less specific. Thus, a decrease in the lymphocyte count may be the most important monitoring for routine biochemical tests ^[1,6]^. The changes of chest CT in this group were similar to those in other COVID-19 studies, and were significantly different from the characteristics of H1N1 pneumonia ^[17]^.

The treatment measures of the patients in this group were mainly in accordance with the protocol ^[8,9]^. Although the patients were mainly mild and moderate, the treatment was complicated due to the particularity of the epidemic ^[18,19]^. Two-thirds of the patients were given antibiotics, although there was not enough evidence of bacterial infection. Although no specific antiviral drugs were recommended, the patients in this group were routinely given antiviral drugs, and two-thirds were given two antiviral drugs and one-third were given three antiviral drugs. The effects of thymosin, glucocorticoid, albumin, and immunoglobulin on COVID-19 need to be further investigated, especially in mild and moderate patients. Many studies had demonstrated the important role of TCM in inhibiting coronavirus. ^[20,21,22]^

Although the patients in this study were mainly mild and moderate, there was still a significant difference in the duration from onset to release form quarantine. The most important basis for release from quarantine is the persistence of negative nucleic acid test results, so the duration from onset to release form quarantine reflects the time it takes the patient to release the virus from the respiratory tract. The average time from onset to release form quarantine was 20 days. Since the patient could only be released from quarantine after three consecutive negative nucleic acid test results, and each test interval was 24 hours, and the incubation period reported in previous literature was considered ^[1]^, the average duration of virus release in this study should be similar to that of professor Cao Bin’s study.^[23]^

Some items in the ≤ 20-day group and the > 20-day group were significantly different, which may be the reason why the quarantine cannot be released for a long time. The age of the > 20-day group was older and the time from onset to admission was later, suggesting that although there was no specific antiviral drug for COVID-19, systematic supportive treatment after admission could improve outcomes, even in mild and moderate patients. There was no difference in PSI score, chronic underlying disease, epidemiological history on admission between the two groups, possibly due to the small number of cases or mild illness. Symptoms on admission, 7 asymptomatic patients screened recovered quickly, which may be related to the viral load and individual differences. Symptomatic patients, fatigue and pharyngeal pain were more obvious in patients in the > 20-day group, for unknown reasons. In terms of routine blood biochemical examination and pulmonary imaging, although the abnormal proportion of individual indicators in the >20-day group was higher than that in the ≤20-day group, the number of samples was not large enough to have sufficient clinical significance. There was no significant difference in the number of patients between the two groups, which was related to the fact that almost all of the patients were mild and moderate, while one critically ill patient was quickly released from quarantine. These clinical results may suggest that there is a cross relationship between the sustained positive nucleic acid of novel coronavirus in respiratory tract specimens and the severity of the disease ^[1,23,24]^, but it is not a linear relationship, and the reasons for the sustained positive nucleic acid are complex. The > 20-day group used more drugs, which may be related to the eagerness to make the patient’s nucleic acid negative.

In particular, there was no significant improvement in the white blood cell and lymphocyte counts at the time of release from quarantine and at the time of admission in either the ≤20 days group or the > 20 days group. The reasons need to be further studied.

The number of cases in this study is not large and has obvious regional characteristics, and the disease is mainly mild and moderate, which cannot represent the characteristics of a large number of patients in a large geographical range. It is also not representative of severe and critical patients.

## Conclusion

Most of the COVID-19 cases in Liaocheng city are mild and moderate. The main cause of infection is exposure to a confirmed patient or the workplace of a confirmed patient. The main clinical symptoms are cough, fever and fatigue, but shortness of breath, sore throat and gastrointestinal symptoms are also common. Chest CT scan showing pneumonia and reduced lymphocyte count are the most important adjunctive examination. The duration released from quarantine was related to age, the length of time from onset to admission, the presence or absence of symptoms, and was not related to mild or normal type. There was no significant improvement in white blood cell and lymphocyte counts at the time of release from quarantine compared to the time of admission.

## Data Availability

All data referred to in the manuscript is available.

---

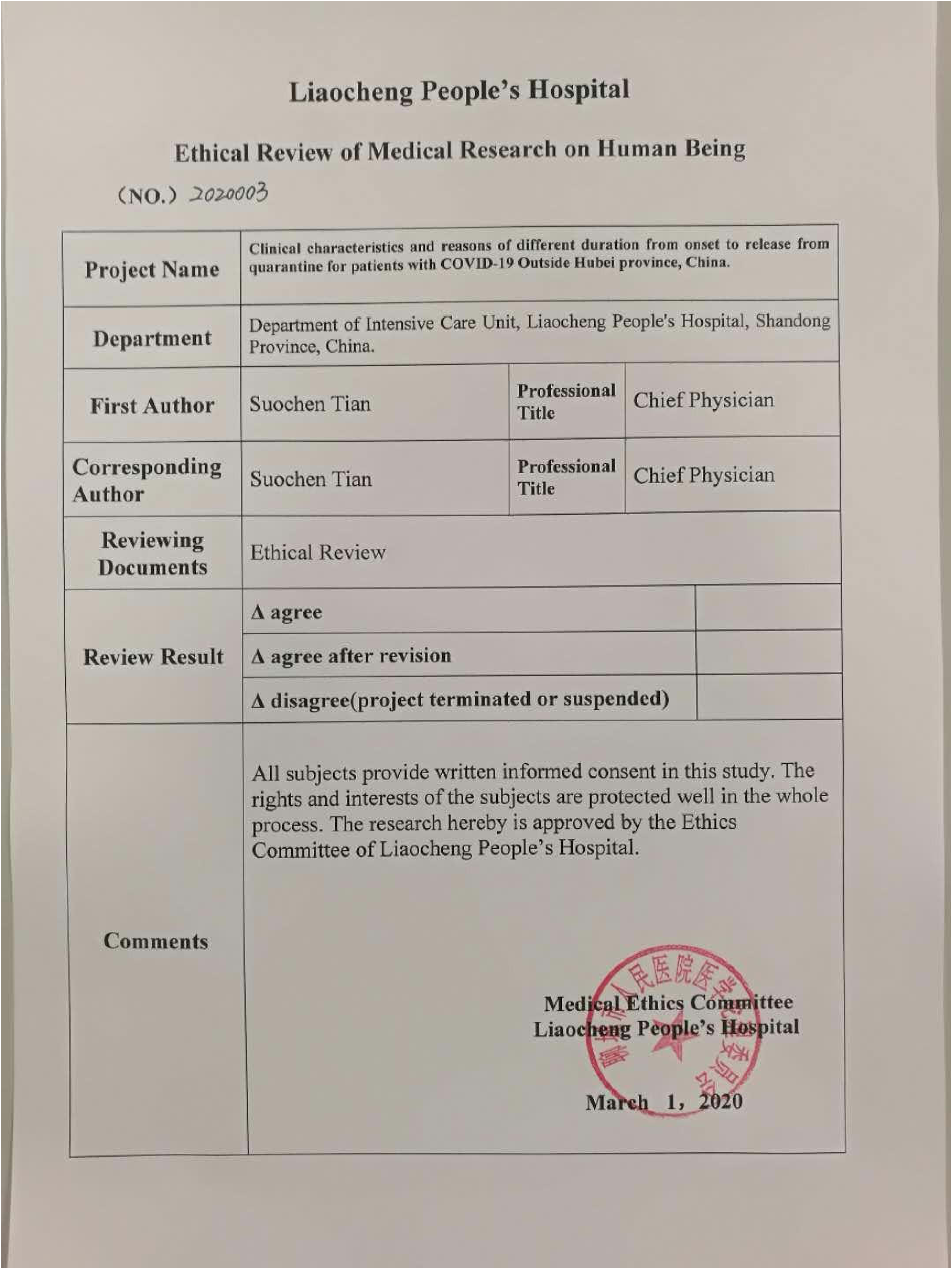

